# Public communication and outreach by mosquito programs in the United States

**DOI:** 10.1101/2024.09.17.24313841

**Authors:** Nicole Thomas, Jo Anne G. Balanay, Sachiyo Shearman, Stephanie L. Richards

**Affiliations:** Department of Health Education and Promotion, East Carolina University, Greenville, North Carolina, United States of America; School of Communication, East Carolina University, Greenville, North Carolina, United States of America

## Abstract

Blood feeding female mosquitoes cause itchy welts and can transmit pathogens that cause diseases such as chikungunya, malaria, West Nile encephalitis, and Zika. Mosquito control programs conduct mosquito, pathogen, and epidemiological surveillance, carry out source reduction, treat mosquito habitats with larvicides or adulticides, and disseminate information to the public. Here, 100 organizations (e.g., private/public mosquito control programs, national professional mosquito/pest control associations) in the United States were asked to complete a survey (N=39 respondents) about their public communication and outreach efforts. Results indicate most programs (N=27, 69%) have dedicated personnel for public communication. A checklist was constructed to compare communication strategies between a subset of program websites and Facebook pages. Recommendations for improving public communication and outreach strategies for mosquito control programs are discussed.

## Introduction

Mosquitoes (Order Diptera, Family Culicidae) transmit pathogens that impact public and veterinary health (e.g., yellow fever [virus], dengue [virus], West Nile [virus], chikungunya [virus], filariasis [worm], and malaria [protozoan]) [1]. Mosquito control programs (MCP) protect public health from mosquito-related issues using methods such as community education, source reduction, bed nets, larvicides, adulticides, biological control, and release of sterilized or genetically altered mosquitoes [1, 2, 3]. Surveillance informs operational decisions of MCP and helps programs evaluate intervention efforts [4].

Social media is one of the largest growing forms of community outreach in the United States (US) and 93% of adults use the internet [5]. Facebook is the most used social media platform (68% of internet users) [6, 7]. In some cases, people prefer social media platforms over mainstream outlets as a news source due to a stronger sense of connection by direct association or perceived closeness [8, 9]. Program utilization of different media platforms including social media, podcasts, television, and websites could increase public knowledge and awareness of mosquito-borne diseases and methods for prevention. Internet-based sources include micro-blog posts (e.g., Facebook and X [formerly Twitter]), web encyclopedias, search queries, and other forms of social media [10]. These sources are especially helpful in areas where public health agencies are limited such as developing countries [10]. In some cases, public health agencies deal with public distrust of government officials to provide safe solutions (e.g., yellow fever vaccine) [9]. Media platforms are readily available to public health agencies; however, lack of real-time information distributed through these methods can lead to misinformation [11].

Through investigation and analysis of social media and other outreach methods used by county/state/federal agencies, private pest control companies, and professional public health pest control associations, public education about mosquito-borne disease and mosquito control could be streamlined and improved. There has not yet been another comparative analysis of such entities. Improvement of outreach effort can facilitate public communication by MCP and combat misinformation. Analysis of keywords in public social media messages can help identify early indicators to integrate infodemiology (i.e., online information about human behavior) and/or infoveillance (i.e., surveillance of online information) into public communication campaigns [10, 12]. Exploration of public search and commenting queries on social media outlets can show areas of rising public concern related to potential outbreaks. Google search demands have been previously analyzed for a flea-related disease outbreak of plague [13] and similar methods could be implemented for mosquito-borne diseases. Google Trends (GT) can track internet search history for a specified range of time using term and topic searches [13]. Spatiotemporal search parameters allowed GT to monitor relationships between public internet searches and confirmed plague cases in the region of interest [13].

Investigation into methods used by the public when conducting illness queries can be a starting point for surveillance data. Some of the public may post on Facebook to obtain opinions, while others may use Google or YouTube to search for information [8]. Some individuals search government health agency websites for information, and others utilize social media for broadcasting on X about symptoms [8]. The public generally seeks information when health risk uncertainty rises and may engage in popular social platforms (i.e., Reddit) for posting questions soliciting public response [8]. Emphasis placed on illness symptom searches or posts is “syndromic surveillance” [14]. On platforms such as X, an individual can post symptoms and their geo-location data is recorded [14]. By highlighting key words and emojis, symptoms can be tracked, and outbreak trends confirmed to evaluate risk [14]. Countries, such as the US, United Kingdom, and China track influenza trends by comparing social media trends with confirmed outbreaks [14]. The same study showed that, where locations were tracked, relevant tweets were positively correlated with observed public health data. On a larger scale, predictions of epidemics could be made from social media data which could help track transmission of vector-borne, food-borne, and other illnesses [10].

One obstacle facing public health educational campaigns is determining the best method(s) for disseminating information including evaluation of: 1) information source, 2) messaging, 3) audience, and 4) method of information delivery (e.g., news, blog, social media) [6, 15]. Hence, the evaluation of outreach methods used by public and private health agencies would be beneficial [6]. To combat misinformation, public health agencies should effectively distribute accurate information to the public [15]. The internet presence of public health agencies should engage the public through resources such as social media [15]. Public and private MCP must also engage with the public via social and traditional types of media and advertising must be balanced with reliable information to maintain credibility [8].

Public interaction strategies should provide up-to-date information and enhance engagement with social media posts [15]. Knowledge of diverse disciplines (e.g., business, psychology, marketing) can enhance public health messaging [6]. Scientists should succinctly relay findings to be received by audiences of different backgrounds and cultures to increase the likelihood of reading and sharing [16]. Improved marketing tools can help structure messaging to increase positive perception of information [6]. Public health organizations often disseminate information via academic journals and academic conferences [6]. Higher value is placed on information received by people with whom the recipient has an established relationship or familiarity [9]. Therefore, increasing internet and social media presence is vital for public health organizations to help establish themselves as consistent and knowledgeable sources of information to the community [15].

Daily news reporting is related to public internet searches; hence, these sources can be paired to streamline and improve communication [17]. This relationship was observed during the 2016 Zika virus outbreak and showed an avenue of information sharing to further mosquito control education [17]. Increasing public awareness of public health pests and mosquito-borne diseases can be a joint effort between different cooperating agencies and industries. Risk communication, messaging, and other forms of outreach can be improved when multiple specialties provide input that considers audience diversity. Through utilization of market research and business strategies, public health agencies can incorporate cost-effective communication methods to disseminate information [6, 16]. Social media outlets provide an opportunity for cost-conscious public health messaging [11]. Risk communication plans should be in place and implemented before an outbreak. Proactive evaluation of current methods of communicating public health pest information would inform policy recommendations to improve accessibility. Consequently, the objectives of this study were to: 1) Identify and describe current public communication and outreach methods for disseminating information on mosquito-borne disease awareness and prevention by county/state/federal agencies, private vector/pest control companies, and professional public health pest control associations, and 2) Analyze communication and outreach methods regarding mosquito-borne disease to provide recommendations on improving future communication.

## Materials and Methods

### Survey on Public Communication

Mosquito control programs vary in size and scope across the US; hence, differences in public communication methods were expected between programs. To assess and compare MCP communication efforts, a 23-question survey was developed in Qualtrics and administered by email to 100 MCP and other mosquito-related organizations across the US from March 7-22, 2024 (Appendix). The survey was approved by the East Carolina University Medical Center Institutional Review Board (UMCIRB# 24-000301). Participants provided written consent prior to completing the survey. Questions included topics such as geographic location, funding level, mosquito-borne diseases of concern, frequency/ types of communication utilized, status of communication staff, and availability of funding for communication.

The following types of MCP were contacted: state agency (SA), federal agency (FA), private pest control agency (PR), public pest control (PU), and professional public health pest control associations (PA). Programs in the following US regions were contacted: 1) North-Atlantic (N=10), 2) Mid-Atlantic (N=21), 3) South-Atlantic (N=10), 4) North-Central (N=15), 5) West-Central (N=10), 6) South Central (N=13), 7) North-Pacific (N=10), 8) South-Pacific/Pacific (N=10). One federal agency (no headquarter location provided) was also included (Table 1).

**Table 1.**
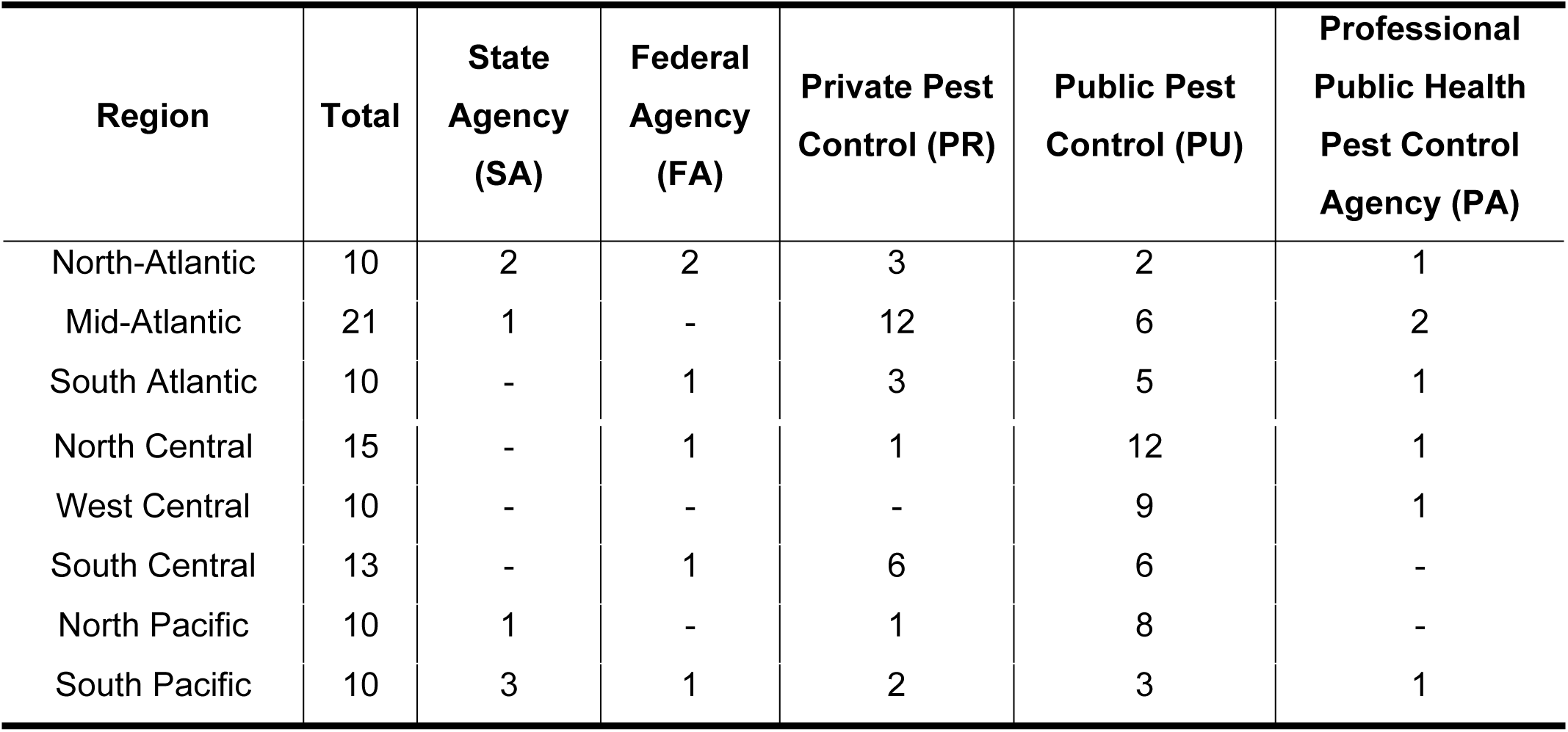
Regions and agency types for 100 programs contacted.

| Region | Total | State Agency (SA) | Federal Agency (FA) | Private Pest Control (PR) | Public Pest Control (PU) | Professional Public Health Pest Control Agency (PA) |
| --- | --- | --- | --- | --- | --- | --- |
| North-Atlantic | 10 | 2 | 2 | 3 | 2 | 1 |
| Mid-Atlantic | 21 | 1 | - | 12 | 6 | 2 |
| South Atlantic | 10 | - | 1 | 3 | 5 | 1 |
| North Central | 15 | - | 1 | 1 | 12 | 1 |
| West Central | 10 | - | - | - | 9 | 1 |
| South Central | 13 | - | 1 | 6 | 6 | - |
| North Pacific | 10 | 1 | - | 1 | 8 | - |
| South Pacific | 10 | 3 | 1 | 2 | 3 | 1 |

### Website and Social Media Analysis

A media content checklist was developed to evaluate a subset of respondent organizations for items such as: Feedback provided to the public, mosquito-borne disease awareness and prevention information, and messaging delivery systems/methods. This checklist allowed for comparison of websites with social media resources. The checklist assessed: 1) program purpose, 2) mosquito-borne disease (e.g., diseases of concern, numbers of cases/illness/death), 3) prevention strategies, and 4) information delivery methods (e.g., positive/negative messaging [benefits vs. risks], presence/absence of images, ability of audience to provide feedback). The checklist was completed for a subset of five websites and related Facebook pages including private pest control programs (this group was underrepresented in the survey respondents), municipal mosquito control, and statewide non-profit mosquito/pest association. These five organizations were randomly selected based on the presence of a website and Facebook page. Facebook pages were evaluated for posts made between January 1 - December 31, 2023.

### Data Analysis

A total of 39 (39% response rate) complete surveys were received in Qualtrics and data were analyzed with Statistical Package for the Social Sciences (SPSS 29, IBM, Armonk, New York). Chi-square (*P*<0.05) was used to assess the association between reported funding and personnel dedicated to risk communication.

## Results

Some respondents skipped questions; therefore, total numbers may vary in figures. No responses were received from private pest control programs. Respondents were from the following US regions and states: North-Atlantic (N=2, 5%; New York), Mid-Atlantic (N=3, 8%; North Carolina, Virginia), South-Atlantic (N=5, 13%; Georgia, South Carolina), North-Central (N=13, 33%; Illinois, Michigan, Missouri, Ohio, Wisconsin, Native American reservation between Wisconsin and Michigan), West-Central (N=7, 18%; Colorado, Utah, Wyoming), South Central (N=2, 5%; Louisiana), North-Pacific (N=1, 3%; Washington), and South-Pacific (N=6, 15%; Arizona, California).

Respondent programs included: district (N=14; 36%) county environmental/public health (N=10; 26%), other (N=8; 21%)(e.g., Indian Health Service, US Environmental Protection Agency, and county mosquito programs through public works), state health (N=2; 5%), large county (N=2; 5%), state/regional professional association (N=1; 3%), federal health (N=1; 3%), and city public works (N=1; 3%) (Fig. 1). Programs reported an average of $900,000 in total annual program funding, ranging between $0 to $20,000,000.

**Fig. 1.**
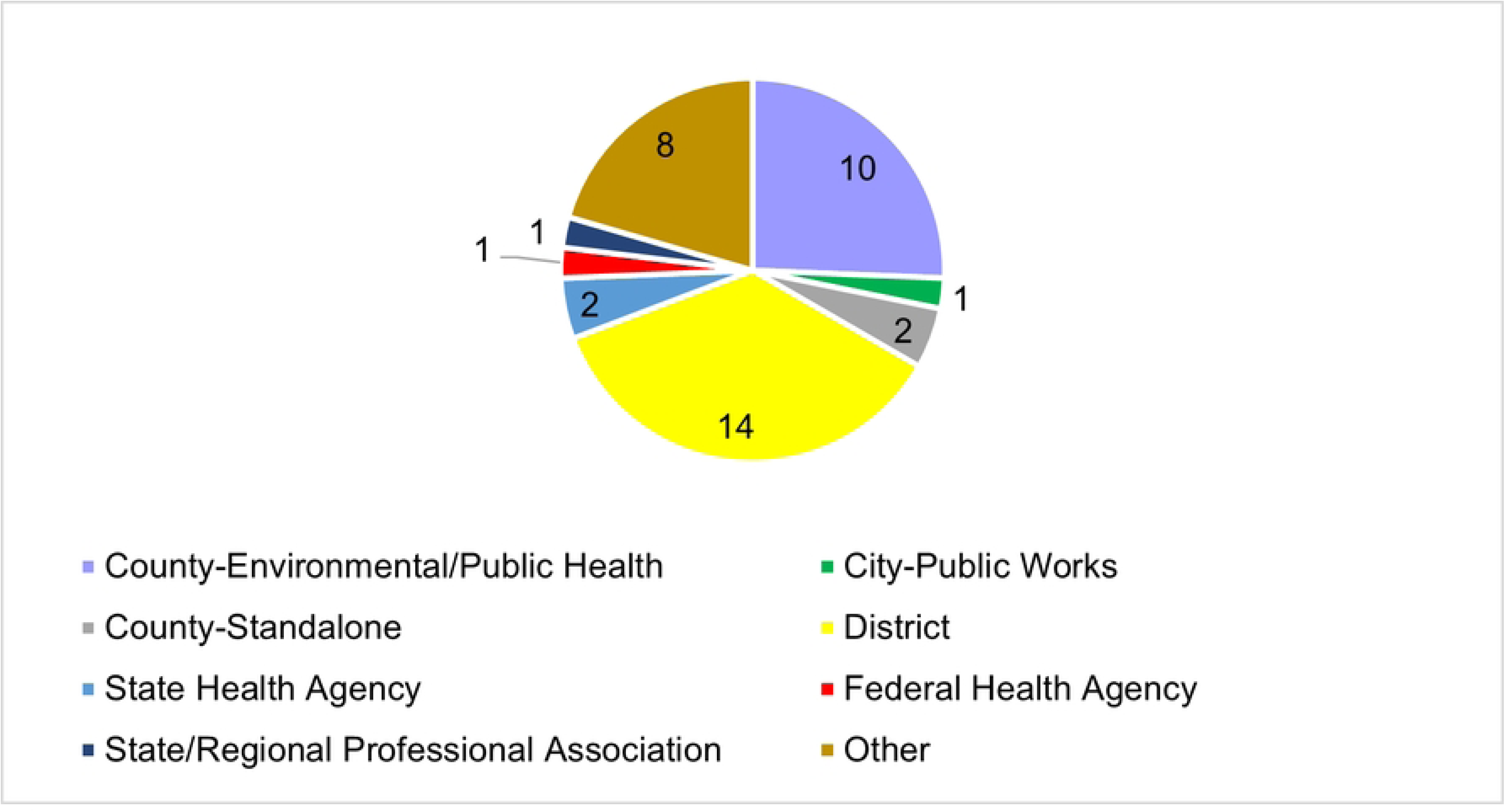
Program types of survey respondents.

Some respondents (N=20; 51%) indicated their program covered a larger region than where they were based, while others (N=19; 49%) stated their program covered only the region in which they were located. Programs covering a larger region indicated coverage areas included other municipalities, counties, and multiple states.

Respondents (scale of 1-10; 1=weak, 10=strong) indicated an emphasis of 7.7 ± 2.4 (mean ± standard deviation) on information-sharing regarding the specific diseases of prevalence in their area. Examples of reasons for low ratings included, “we support other entities in doing this but do not do so ourselves”, “we have no one dedicated to public outreach and education”, and “we can do better”. None of the respondents select a value of 1. Low emphasis ratings (i.e., 2-4) comprised 18% of the total responses. Medium emphasis ratings (i.e., 5-7) were 8% of responses. Most (74%) indicated a high emphasis (i.e., 8-10) on information sharing.

Most respondents (N=27; 82%) indicated having a dedicated public communication outreach division for dissemination of information about vector-borne disease and/or mosquito control. Five (15%) respondents indicated not having a public outreach division and one (3%) was unsure. However, 34% (N=11) of respondents indicated their agency had funding dedicated to public communication, 53% (N=17) did not have funding, and 13% (N=4) were unsure. Chi-square analysis showed dedicated communication funding was significantly associated with dedicated communication personnel (*Χ*^2^=43.04; *P*<0.01).

Communication methods utilized by respondents included: 1) in-person/phone outreach (N=29, 26%), 2) website (N=27, 24%), 3) printed brochures/advertising (N=26, 23%), 4) social media (N=19, 17%), 5) virtual advertising (N=8, 7%), and 6) other (N=4, 4%)(i.e., press releases and news broadcasts) (Fig. 2).

**Fig. 2.**
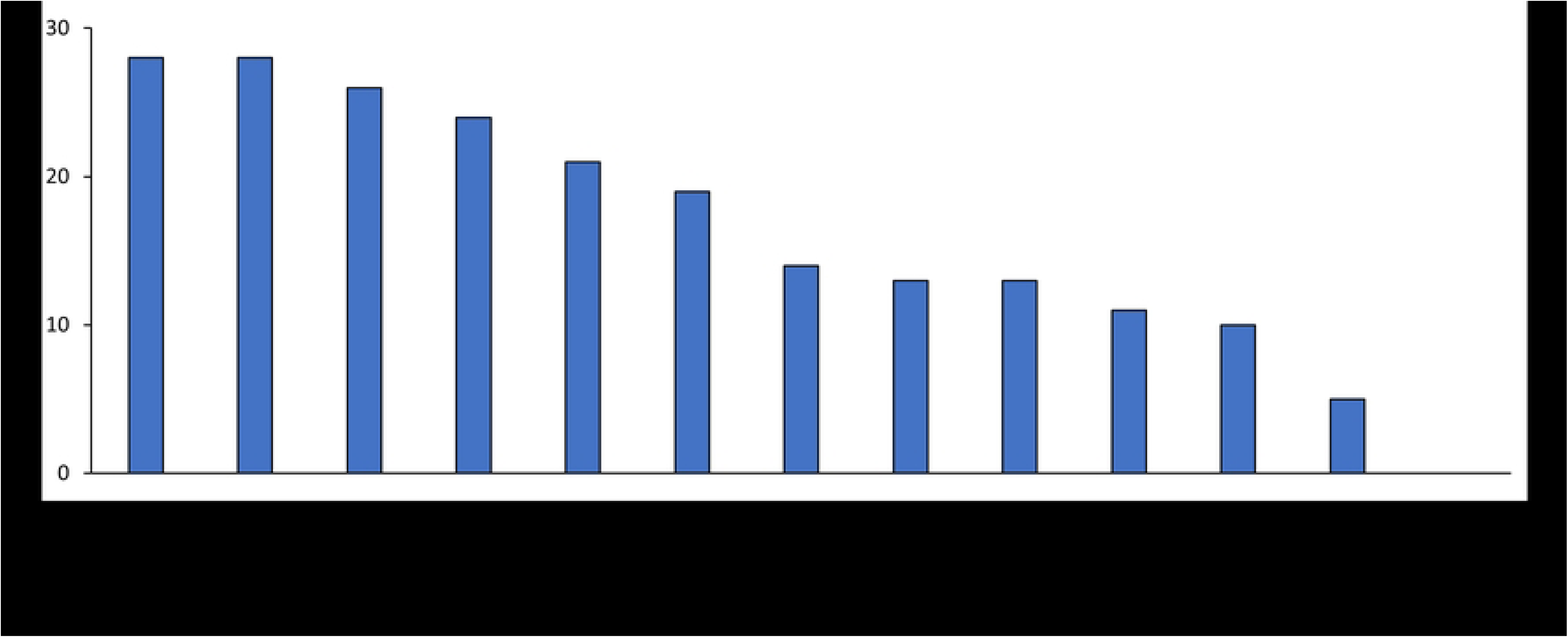
Methods of public communication used by respondents. Insert figure here.

The primary communication methods indicated by respondents included: 1) response to citizen pest complaints via site visits (N=28, 13%) and phone (N=28, 13%), 2) website (N=26, 12%), 3) brochures (N=24, 11%), 4) email (N=21, 10%), 5) booth at local fair (N=19, 9%), 6) Facebook (N=14, 7%), 7) Instagram (N=13, 6%), 8) workshop/conference (N=13, 6%), 9) visits to schools for education (N=11, 5%), and 10) local media outlet advertisements (N=10, 5%) (Figure 2). When programs were asked about types of online communication, most (N= 26, 48%) was informational, followed by risk communication for an impending health concern (N=23, 43%), or other (N=3, 6%) (e.g., promotion of services or press releases), and some programs selected ‘other’, indicating they did not use online communication (N=2, 3%).

Images (N=26; 93%) were frequently used in online public communication messages, followed by web links (N=25, 89%), text (N=23, 82%), infographics (N=19, 68%), and alteration of fonts (N=18, 64%). One respondent (4%) also reported using quick response (QR) Codes. Respondents indicated that positive framework/phrasing was their primary delivery method (N=19, 90%) while negative framework (N=6, 29%) was used less frequently. In some cases, audiences could provide feedback (N=8, 38%) and paid advertisements (N=4,19%) were also utilized. When customizing information for social media, programs shorten messaging (N=14, 50%), modify messages for the target audience (N=11, 39%), or make no modifications to messaging (N=10, 36%). Some respondents indicated restricting public comments on social media (N=4, 14%) and/or selected ‘other’ (i.e., messaging handled by a public information officer or is posted on their website [N=2, 7%]).

Respondents indicated communicating with the public always (N=17, 57%) or most of the time (N=9, 30%) and most (N=19, 63%) never communicated with agricultural industries. Most programs communicated with beekeepers sometimes (N=10, 33%), most of the time (N=7, 23%), or never (N=7, 23%). Most programs communicated with health professionals sometimes (N=10, 40%) or never (N=7, 23%). Programs not communicating with the public listed these barriers: lack of time (N=11, 25%), lack of communication personnel (N=11, 25%), lack of funding (N=8, 18%), lack of social media expertise (N=8, 18%), and/or perceived lack of public interest (N=2, 5%).

A checklist was completed to analyze program websites and Facebook pages for five private pest control programs over a one-year period. An additional evaluation was carried out on the most active Facebook post. Media content analysis included private pest control companies (N=3, 60%), city mosquito control (N=1, 20%), and a statewide non-profit mosquito/pest association (N=1, 20%). Programs showed information about mosquito borne diseases (N=3, 60%) where diseases of concern (N=2, 40%) and total number of illness/deaths (N=1, 20%) were listed. Most (N=3, 60%) did not include prevention strategies to protect against mosquito-borne illness. Most Facebook pages included prevention strategies (N=4, 80%) that focused on treatment options and sales and allowed the public to leave comments. All programs provided images on both their website and Facebook page, including mosquitoes/vectors (N=4, 80%), treatments (N=3, 60%), positive people (N=2, 40%), and the environment (N=1, 20%).

The Facebook analysis showed that all programs (N=5, 100%) provided information in a positive framework whereas one program (20%) also used negative framework in their messaging. All websites (N=5, 100%) showed a statement of purpose and links to additional information on external websites. However, 60% (N=3) of Facebook pages did not include a statement of purpose. The average number of agency posts on all five Facebook pages during the study period was (mean ± standard deviation) 58.6 ± 47.1 and 51.7 ± 85.3 images were shared via Facebook posts. When the most popular post (21 likes/comments/shares ± 24.5) on each program’s Facebook page was analyzed, 57% (N=4) of the posts focused on public participation (e.g., community meeting), while others included information about disease prevention and awareness.

## Discussion

Vector control agencies communicate with the public in a variety of ways including, but not limited to television, radio, newspaper, website, and social media [18]. Social media showed an increase in communication frequency between the public and health organizations during the Zika epidemic in 2015-2016 [19]. Social media allows individuals not connected to traditional media sources to remain informed [3]. Web links can be provided for users to find additional information and this method is frequently utilized [3]. Environmental health literacy can facilitate risk communication by helping vector control programs address audiences from a variety of backgrounds [20]. This study showed that site visits and phone calls are the most utilized methods of public communication, and that social media are underutilized for reaching a larger audience. Organizations involved in information dissemination on mosquito control and mosquito-borne disease prevention need to give more attention to improving social media use as a potentially efficient and effective communication tool for the public.

Previous studies show low survey participation in states that have mosquito control programs with lower budgets, which may have also limited survey responses here [21]. Limited funding may have also contributed to the lack of survey participation by private pest control companies, as funding efforts are likely weighted towards field personnel and equipment [21]. This should be investigated further. Funding for public communication was related to the presence of dedicated communication personnel, which may imply the importance of financial resources in improving an organization’s communication and outreach program.

### Study Limitations

Although we had a relatively high (39%) survey response rate from a variety of different types of programs across the US, it was difficult to recruit private pest control companies to complete the survey due to lack of direct contact information. Program funding reported by respondents varied widely between programs (range, 0 to $20 million) and budgetary constraints for communication efforts varied, likely due to program size and priorities.

Analysis of a subset of programs’ social media and websites was successful but may be limited due to the willingness of the public to engage with posts [11]. Even if someone positively views a post, they may or may not respond to the post [11]. This study did not consider the number of times a post was viewed but this may be considered if other social media platforms are analyzed.

### Future Studies

Information about public communication methods of private pest control programs should be studied further and compared to public programs. Future studies may consider implementing public communication strategies and assessment of efficacy within different types of audiences (Lindsey, 2022). It would be beneficial for future research to expand the number of organizations whose media content is analyzed through checklists, including county, state, and federal programs.

### Conclusions and Recommendations

Communication personnel existed in most surveyed programs and many respondents use social media as a communication tool. Several organizations prioritize communication about mosquito control and mosquito-borne diseases; however, these efforts are often handled by a public information officer or at a federal rather than regional level. Notably, this study found that the primary reasons for lack of online communication are due to the lack of personnel with social media knowledge and lack of personnel dedicated to communication, hence this deficiency could be improved with training on public communication via social media. Mosquito control programs could consider redistributing funding and/or priorities to increase public risk communication efforts. Posting information about disease prevention and community involvement would likely help programs increase public engagement, awareness, and possibly long-term support for programs.

## Data Availability

Survey data are provided in the manuscript. More detailed (anonymous) survey data can be requested from the corresponding author.

## Acknowledgements

The authors thank the many mosquito control program directors and others that provided valuable feedback for our survey.

## Author Contributions

Conceptualization: Jo Anne G. Balanay, Stephanie Richards, Sachiyo Shearman, Nicole Thomas.

Data curation: Nicole Thomas. Formal analysis: Nicole Thomas.

Investigation: Jo Anne G. Balanay, Stephanie Richards, Sachiyo Shearman, Nicole Thomas. Methodology: Jo Anne G. Balanay, Stephanie Richards, Sachiyo Shearman, Nicole Thomas. Project administration: Jo Anne G. Balanay, Stephanie Richards, Sachiyo Shearman, Nicole Thomas.

Writing – original draft: Nicole Thomas, Stephanie L. Richards.

Writing – review & editing: Jo Anne G. Balanay, Stephanie Richards, Sachiyo Shearman, Nicole Thomas.

## Appendix

### Survey Questions

1. Please list the location information for your mosquito control or other pest/vector-related program: City: ____________________ County: ____________________ State: ____________________
2. Does your program cover a larger region or area (beyond the area of program residence)? If yes, please describe.
  - Yes If yes, please describe: ____________________
  - No
3. Please select the choice which best describes your program type:
  - County mosquito control program within environmental/public health department
  - City mosquito control program within public works department.
  - Large county mosquito control effort
  - Mosquito control district
  - State health agency
  - Federal health agency
  - State or regionally funded professional association
  - National professional association
  - Non-profit mosquito/ pest association
  - Private pest control company
  - Other: ____________________. Please describe.
4. On the scale of 1 to 10, 1 = low emphasis and 10 = strong emphasis, how much does your program emphasize public communication regarding specific mosquito-borne disease(s) and/or other vector-borne diseases in your region?
5. Please explain why you selected your emphasis rating ____________________
6. What is the approximate total annual budget of your vector control program? ________________________________________________________
7. Please list the approximate percentage of vector control program budget dedicated to each of the following:
  - Public education
  - Mosquito or other pest surveillance
  - Arbovirus surveillance: List arboviruses surveyed: _______________________
  - Mosquito control (e.g., larvicide, adulticide, source reduction)
  - Control of other vectors/pests (Please specify.)
  - Other:
8. From what source does your program receive funding? If funding comes from a range of sources, please list approximate percentage received from each source below.
  - Donations: ______________________
  - City/County Government: ______________________
  - State Government: ______________________
  - Federal Government: ______________________
  - Federal, State, or Local Government Grant (Please Specify): ______________________
  - Other: ______________________
  - Unsure/Don’t know
9. Does your organization or program have a division, group, and/or individual personnel dedicated or partially dedicated to public communication/outreach?
  - Yes If yes, please specify: ______________________________________
  - No
  - Unsure/ Don’t Know
10. Does your program have dedicated funding for public communication efforts about vector-related issues (e.g. mosquito-borne or other vector-borne diseases, mosquito control treatments, etc.)?
  - Yes
    - If yes, approximately how much funding is dedicated to communication? Please describe.
  - No
  - Unsure/ Don’t Know
11. What are the primary methods of public communication utilized by your program? Please assign the items below a number, based on your program’s frequency of use, where 1 is the most frequently utilized. Please list 0 for items that do not apply.:
  - Facebook
  - Instagram
  - Email
  - Website
  - Response to citizen pest complaints via site visit and/or in person consultation
  - Response to citizen pest complaints via phone
  - Workshop/ Conference
  - Booth at local fair
  - Visit to schools for education
  - Brochures
  - Local media outlet advertisements (e.g., regional news broadcasts, local newspapers, etc.). Please list and describe.
  - My program does not conduct public communication.
  - Other: _________________. Please describe.
12. If your program conducts public communication, what types of messaging are used? Please assign the items below a number, based on your program’s frequency of use, where 1 is the most frequently utilized. Please list 0 for items that do not apply. Beside each applicable category, please specify if this communication is virtual or in-person:
  - Informational (related to a notable subject or specific pest)
  - Risk communication (notifying the public of an impending health risk/concern facing members of the community)
  - My program does not conduct public communication.
  - Other: __________________. Please describe.
13. If your program conducts online public communication, approximately how frequently does your program update communication messaging? Select one and please specify if this communication is virtual or in-person:
  - Once or more per week.
  - Biweekly
  - Monthly
  - Seasonally
  - Bi-annually
  - Annually
  - As needed if there is a disease outbreak
  - As needed to provide details about mosquito control treatments
  - My program does not conduct online public communication.
  - Other: __________________. Please describe.
14. Please describe program plans (if any) to initiate or further develop mosquito/pest communication. ______________________________________________________________________________
15. If your program does not conduct public communication, please describe barrier(s) that may impact this. Choose all that apply:
  - Lack of time
  - Lack of funding
  - Lack of/limited personnel dedicated to communication.
  - Lack of personnel expertise in social media or other online communication skills.
  - Perceived lack of public interest.
  - Other: ____________________. Please describe.
16. What communication platforms are utilized by your program?
  - In-person/Phone Outreach
  - Printed Brochures/Advertisement
  - Virtual Advertisement
  - Program Website
  - Social Media
  - Other – Please specify: _____________________
17. What is the approximate percentage of communication that is Virtual or Online versus In-Person/Phone Calls?
  - Virtual or Online: __________________
  - In-Person/ Phone: __________________
18. How is communication modified across media platforms used by your program? Select all that apply:
  - Abbreviated message for social media
  - Comments restricted on social media
  - Messages modified to reflect target audience
  - Other – please explain.
  - No modifications
19. What items are included in your public outreach messages? Select all that apply.
  - Test
  - Infographics
  - Images
  - Font alteration (e.g. change of colors, increasing boldness, including italics)
  - Using web links
  - Other – please specify. _______________________________
20. Please describe your communication methods regarding delivery. Select all that apply.
  - Positive Framework/Phrasing
  - Negative Framework/Phrasing
  - Paid advertisements. Please specify (e.g. billboards, vehicle decals, virtual ads)
  - Allowing audience feedback – please specify if this is via survey, phone calls, or public comments
21. How often do you target the following audiences with your mosquito/vector-related information?
  - General public
    - Never, Rarely, Sometimes, Often, Always
  - Health professionals
    - Never, Rarely, Sometimes, Often, Always
  - At risk populations (e.g., immunocompromised, elderly people sensitive to insecticides)
    - Never, Rarely, Sometimes, Often, Always
  - Beekeepers
    - Never, Rarely, Sometimes, Often, Always
  - Agricultural Industries
    - Never, Rarely, Sometimes, Often, Always
  - Other (Please describe.): ____________________________
22. Please provide any additional feedback on communication here: ___________________________________

